# Hospital-onset bacteraemia and fungaemia as a novel automated surveillance indicator: results from four European university hospitals

**DOI:** 10.1101/2024.09.16.24310433

**Authors:** Seven J.S. Aghdassi, Suzanne D. van der Werff, Gaud Catho, Manon Brekelmans, Luis A. Peña Diaz, Niccolò Buetti, Ferenc D. Rüther, Daniel Dinis Teixeira, Daniel Sjöholm, Pontus Nauclér, Michael Behnke, Maaike S.M. van Mourik the PRAISE-HOB working group

**Author notes:** These authors contributed equally to this work and share first/last authorship (Seven J.S. Aghdassi, Suzanne D. van der Werff, Michael Behnke, Maaike S.M. van Mourik). The members of the network are listed under Collaborators. **Corresponding author:** Suzanne D. van der Werff, Department of Medicine Solna, Division of Infectious Diseases, Karolinska Institutet, Stockholm, Sweden.

## Abstract

**Background:** Conventional manual surveillance of healthcare-associated infections is labour-intensive and therefore often restricted to areas with high-risk patients. Fully automated surveillance of hospital-onset bacteraemia and fungaemia (HOB) may facilitate hospital-wide surveillance.

**Aim:** To develop an algorithm and minimal dataset (MDS) required for automated surveillance of HOB and apply it to real-life routine data in four European hospitals.

**Methods:** Through consensus discussion a HOB definition with MDS suitable for automated surveillance was developed and applied in a retrospective multicentre observational study including all admitted adult patients (2018-2022). HOB was defined as a positive blood culture with a recognised pathogen two or more days after hospital admission. For common commensals, two blood cultures with the same commensal within two days were required. Annual HOB rates were calculated per 1,000 patient days for the hospital and for intensive care units (ICU) and non-ICU.

**Results:** HOB rates were comparable between the four hospitals (1.0 to 2.2 per 1,000 patient days). HOB rates were substantially higher in ICU than non-ICU across the four hospitals, and HOB with common commensals accounted for 14.8-28.2% of all HOB. HOB rates per 1,000 patient days were rather consistent over time, but were higher in 2020 and 2021. HOB caused by Staphylococcus aureus accounted for 8.4-16.0% of all HOB.

**Conclusion:** Automated HOB surveillance using a common definition was feasible and reproducible across four European hospitals. Future studies should investigate clinical relevance and preventability of HOB, and focus on strategies to make the automated HOB metric an actionable infection control tool.

## Background

Surveillance of healthcare-associated infections (HAI) is a cornerstone of infection prevention and control (IPC) and an effective means to reduce the incidence of HAI (1-3). Current surveillance systems rely primarily on manual chart review to detect infections, which is resource-intensive, time-consuming, and prone to subjective interpretation (4). Automated surveillance (AS) systems utilise algorithms to complement or replace certain steps of manual surveillance by analysing routine care data that are automatically extracted from electronic health records (EHR). AS systems offer potentials to obtain surveillance data in a timelier manner, to increase standardisation and interpretability of results, and once set-up, to be less time-consuming (5-7).

Currently, AS systems are mostly limited to research settings and single institution and hence vary considerably in terms of methods (7). As a result, there is a great need for HAI definitions and targets suitable for AS, reliable detection algorithms, and data structures that promote interoperability of medical data across different institutions. The PRAISE (Providing a Roadmap for Automated Infection Surveillance in Europe) network aims to provide methods and resources for facilitating the transition of AS from the research setting to large-scale implementation (8, 9). Multi-institutional development of AS methods will be crucial to maintain comparability of surveillance outcomes, one of the essential aspects of surveillance.

Hospital-onset bacteraemia and fungaemia (HOB) has been suggested as a novel surveillance and quality indicator and has been considered a suitable primary target for large-scale AS implementation, given the relative simplicity of the case definition with a focus on microbiological results from blood cultures (10). Compared to conventional manual surveillance of central line-associated bloodstream infections (CLABSI), automated HOB surveillance may enable coverage of the entire in-patient populations (i.e., surveillance of all admitted patients not just selected populations) and an increased scope of the surveillance target, by also capturing bloodstream infections (BSI) not associated with intravascular catheters (11). While the potential of HOB surveillance has been discussed in various recent publications (11-13), a detailed HOB definition and method that is suitable for fully automated surveillance, has not been published to date.

In this article, we report the collaborative development of an automated HOB surveillance algorithm within the PRAISE network and its application to retrospective data from four European university hospitals. We aim to illustrate potential applications of HOB surveillance to guide quality improvement and identify questions for future research.

## Methods

### Development of PRAISE-consensus HOB definition

The HOB definition and minimal dataset (MDS) were developed through iterative discussion between members within the PRAISE network. The PRAISE network was formed in 2019 with the ambition to support the development and implementation of automated surveillance in Europe (8). Since 2021, a dedicated HOB working group has focused on developing a HOB definition suitable for large-scale automated surveillance. This PRAISE-HOB working group includes infectious disease and IPC specialists, epidemiologists, and software developers from eight European countries, representing hospitals as well as national reference centres for HAI surveillance.

The following general principles guided the development: feasibility of implementation in all hospitals or hospital networks involved, alignment with existing surveillance methods where possible, and achieving unambiguous definitions of HOB suitable for automated surveillance. No formal consensus methods were used during the development process. Differences were resolved through discussion and, in case of doubt, explorative sensitivity analysis were performed to assess the impact of decisions. The final algorithm is presented in **Appendix 1** and summarised in **Figure 1**.

**Figure 1.**
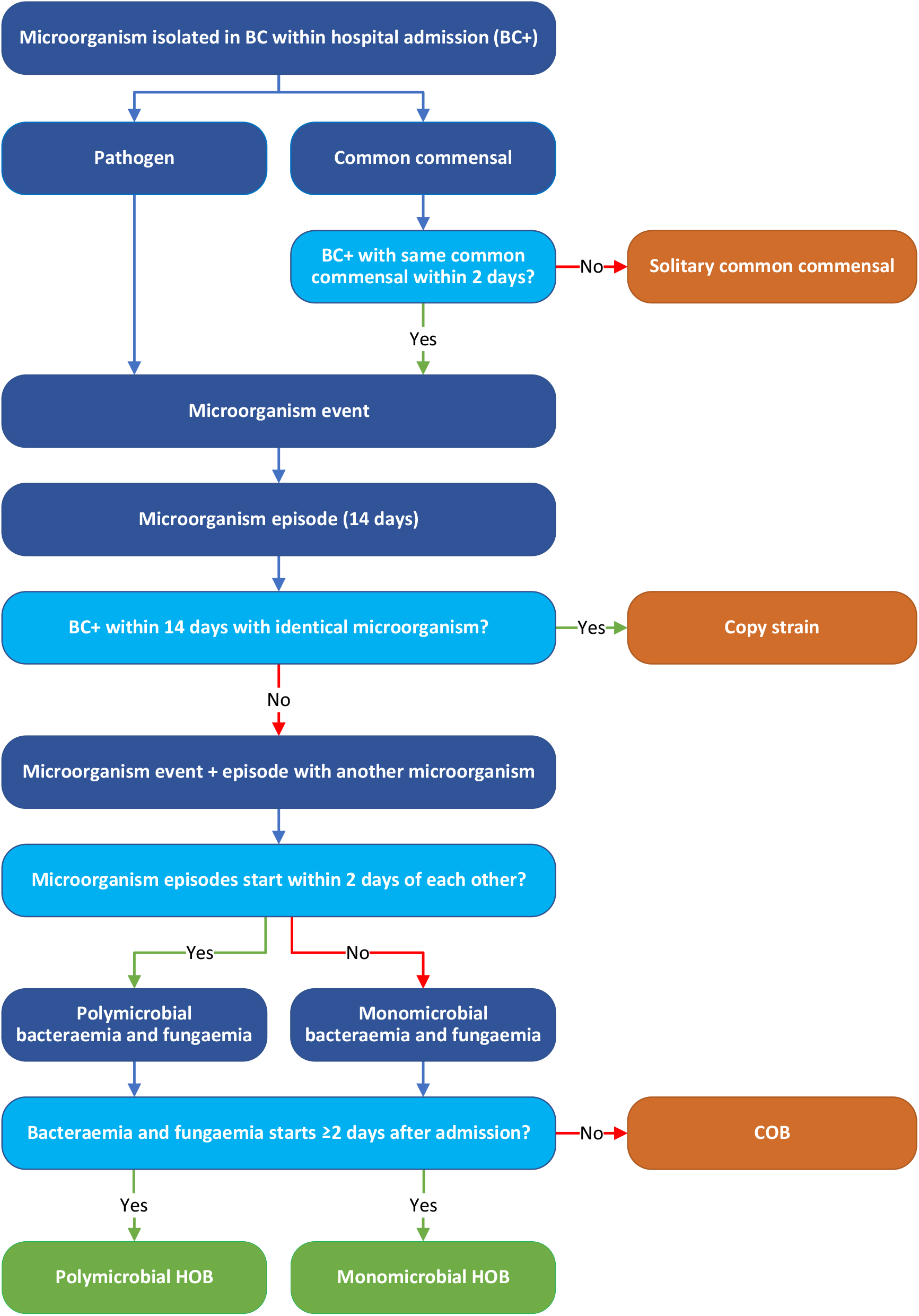
Schematic representation of the HOB algorithm BC = blood culture, BC+ = positive blood culture, COB = community-onset bacteraemia and fungaemia, HOB = hospital-onset bacteraemia and fungaemia. This is a simplified version of the HOB algorithm that is not intended for real-time processing of data, but to visualise how the different concepts relate to each other. For more details, please refer to Appendix 1.

In short, microorganisms identified in blood cultures obtained during hospital admissions are classified as either a pathogen or common commensal as per NHSN (National Healthcare Safety Network) classification of the Centers for Disease Control and Prevention (14). A positive blood culture with a pathogen or two repeated blood cultures with common commensals within two days constitute a *microorganism event*. Solitary common commensals (i.e., common commensals without a confirmation within two days) are disregarded. A microorganism event marks the onset of a *microorganism episode* with a duration of 14 days or until discharge, whichever comes first. During a microorganism episode, the same microorganism in other blood cultures is considered a repeated isolate (copy strain) and does not constitute a new episode. Microorganism episodes starting within 2 days of each other are grouped into *polymicrobial episodes* as they likely represent one clinical event. Finally, the microorganism episodes are classified as hospital-onset, i.e. *HOB*, if they start on day 2 of admission or later (with day 0 being the day of admission). Of note, multiple (overlapping) HOB episodes are possible, for example when different microorganisms are recovered from blood cultures drawn later in time. HOB episodes are attributed to the ward where the patient was two days prior to the start of the episode. Patient days are counted by ward and by groups of wards (e.g., ward type or specialty). In this manuscript, all mentions of bacteraemia reflect bacteraemia and fungaemia.

To support validation and further deployment in different settings, an MDS was defined, specifying the minimally needed data elements for application of the HOB algorithm as well as the required data structure. Multiple rounds of discussion were necessary to define the MDS. **Appendix 1** describes the details of the consensus definition, including examples, a more in-depth depiction of the algorithm, the MDS specification, and details of denominator data calculation.

### Study design and study population

The HOB algorithm was applied in a retrospective cohort study including all patients admitted to one of the participating hospitals during the study period (University Medical Centre Utrecht (Utrecht, the Netherlands), Charité-University Hospital (Berlin, Germany), Karolinska University Hospital (Stockholm, Sweden), and Geneva University Hospitals (Geneva, Switzerland)). University Medical Centre Utrecht has around 1,000 in-patient beds, 27,000 annual in-patient admissions and 170,000 annual patient days. Charité-University Hospital has approximately 3,000 in-patient beds, 120,000 annual in-patient admissions and 800,000 annual patient days. Karolinska University Hospital has around 1,100 in-patient beds, 70,000 in-patient admissions and 370,000 annual patient days. Geneva University Hospital has approximately 2,100 in-patient beds, 50,000 annual in-patient admissions and 700,000 annual patient days. All hospitals are publicly funded tertiary care hospitals (i.e., providing maximum care as per national classification).

The study period varied across hospitals ranging from 2018 to 2022 (see **Table 1**). In this study, paediatric patients, defined as age < 18 or admissions to a paediatric ward, were excluded from the analysis. Moreover, certain wards providing mainly psychiatric, rehabilitation or short-term care, were excluded at the discretion of each hospital. In these cases, denominator data and blood cultures collected from these wards were not included in the denominator or the algorithm. In all hospitals, ethical approval was obtained and informed consent was not needed. If applicable per local regulations, patients who objected to the use of their data for the purpose of research were excluded.

**Table 1.** Description of dataset and rates of HOB per hospital for the entire observation period.

| Parameter | Hospital 1 | Hospital 2 | Hospital 3 | Hospital 4 |
| --- | --- | --- | --- | --- |
| Observation period | 2018-2021 | 2018-2022 | 2018-2020 | 2018-2022 |
| No. of patient days | 454,045 | 3,208,694 | 881,038 | 2,406,577 |
| No. of blood cultures taken | 24,392 | 226,442 | 47,501 | 116,566 |
| No. of blood cultures per 1,000 patient days | 53.7 | 70.6 | 53.9 | 48.4 |
| Blood culture positivity rate (%) | 12.1 | 8.9 | 10.2 | 6.8 |
| No. of HOB episodes | 996 | 6,895 | 1,697 | 2,499 |
| No. of HOB episodes per 1,000 patient days | 2.2 | 2.1 | 1.9 | 1.0 |
| No. of HOB episodes with CC (% of all HOB) | 281 (28.2) | 1,363 (19.8) | 364 (21.4) | 370 (14.8) |
| No. of HOB episodes with CC per 1,000 patient days | 0.6 | 0.4 | 0.4 | 0.2 |
| No. of HOB episodes with pathogen (% of all HOB) | 785 (78.8) | 5,715 (82.9) | 1,405 (82.8) | 2,173 (86.9) |
| No. of HOB episodes with pathogen per 1,000 patient days | 1.7 | 1.8 | 1.6 | 0.9 |
| No. of polymicrobial HOB episodes (% of all HOB) | 159 (16.0) | 849 (12.3) | 226 (13.3) | 303 (12.1) |
| No. of polymicrobial HOB episodes per 1,000 patient days | 0.4 | 0.3 | 0.3 | 0.1 |
| Median no. of in-hospital days until onset of HOB (IQR) | 12 (6-23) | 14 (7-29) | 9 (4-18) | 28 (8-32) |

CC = common commensal; HOB = hospital-onset bacteraemia and fungaemia; IQR = interquartile range; No. = number.
Data depicted in this table pertains only to wards included in the study. Multiple HOB with different microorganisms occurring within a two-day period were counted as one polymicrobial HOB. Since polymicrobial HOB with both pathogen and common commensal were possible, the sum of HOB with pathogen and HOB with common commensal is greater than or equal to the number of all HOB. For counting blood cultures, only those taken from day two onwards were considered to align with HOB counting.

### Algorithm application

All hospitals applied the algorithm locally and only the aggregated outcomes were combined in this manuscript. Data was collected electronically from EHR systems (microbiology results, patient movement data, see **Appendix 1**). To assess whether all algorithms developed by the participating hospitals yielded comparable results, an in-person meeting was organised (July 2023), where only algorithm implementation scripts were exchanged and run on real-life EHR data as well as fictional test data including rare or extreme scenarios (i.e., no sharing of patient data). These datasets were formatted according the MDS. All local algorithms detected the same HOB episodes in both real life and test data.

Algorithms were programmed in different programming languages, depending on the hospital’s expertise and local conventions (SAS enterprise guide, R version 4.1.0, JAVA, or webservice using C# and .Net Core v6).

### Analysis

This manuscript includes an overall presentation of HOB epidemiology in the four European hospitals and illustrates possible applications of HOB surveillance. We present yearly HOB rates, differentiated by pathogen or common commensal, intensive care unit (ICU) or non-ICU, as well as trends over time. Rates are calculated per 1,000 in-patient days. We do not aim to compare rates directly. Instead, we present first insights into the epidemiology of this standardised metric. Furthermore, the frequency of sampling of blood cultures taken on day two after hospital admission or later was depicted to explore the relationship of blood culture frequency with HOB rates. The usability of HOB as a surveillance metric will be further explored by examining the causative microorganisms and drawing careful conclusions from the pathogen distribution. For data presentation, microorganisms were separated into the following groups: anaerobes, coagulase-negative staphylococci, Enterobacterales, enterococci, other Gram-negative rods, *Pseudomonas* species, *Staphylococcus aureus*, streptococci, yeasts and other/unspecified.

## Results

### Cohort description

The four hospitals provided data for variable periods between 2018 and 2022. During the observation period, 2,100 bacteraemia episodes were observed in Hospital 1, of which 996 (47.4%) were HOB. In Hospital 2, 15,096 bacteraemia episodes were recorded, of which 6,895 (45.7%) were HOB. In Hospital 3, 5,411 bacteraemia episodes were recorded, of which 1,697 (31.4%) were HOB. In Hospital 4, there were 6,964 bacteraemia episodes, of which 2,499 (35.8%) were HOB. **Table 1** illustrates the analysed dataset by hospital.

The number of blood cultures per 1,000 patient days ranged between 48.4 (Hospital 4) to 70.6 (Hospital 2). HOB rates per 1,000 patient days were comparable between the four hospitals and ranged from 1.0 to 2.2. The share of HOB with common commensals and pathogens as well as polymicrobial HOB among all HOB was comparable across hospitals. In all four hospitals, the median duration between hospital admission and onset of HOB was over one week, with a range of 9 (Hospital 3) to 28 (Hospital 4) days. Hospital 4 has a longer median and IQR admission time than the other three hospitals because it is organised as a network of several hospitals. Therefore, patients are not considered as readmitted when transferred between different hospitals.

**Table 2** displays the analysed dataset per hospital separately for ICU and non-ICU. In all four hospitals, blood culture sampling frequency and HOB rates per 1,000 patient days were higher in ICU than non-ICU. The proportion of positive blood cultures among all blood cultures taken, however, showed only small differences between ICU and non-ICU. Similarly, the shares of HOB with common commensals and pathogens were comparable between ICU and non-ICU when considered for the individual hospitals.

**Table 2.** Description of dataset and rates of HOB in intensive care units (ICU) and non-ICU per hospital for the entire observation period.

| Parameter | Hospital 1 |  |  |  | Hospital 2 |  |  |  | Hospital 3 |  |  |  | Hospital 4 |  |  |  |
| --- | --- | --- | --- | --- | --- | --- | --- | --- | --- | --- | --- | --- | --- | --- | --- | --- |
|  | ICU |  | Non-ICU |  | ICU |  | Non-ICU |  | ICU |  | Non-ICU |  | ICU |  | Non-ICU |  |
| Observation period | 2018-2021 |  | 2018-2021 |  | 2018-2022 |  | 2018-2022 |  | 2018-2020 |  | 2018-2020 |  | 2018-2022 |  | 2018-2022 |  |
| No. of patient days | 33,335 |  | 420,710 |  | 448,148 |  | 2,760,546 |  | 43,581 |  | 837,457 |  | 50,715 |  | 2,355,862 |  |
| No. of blood cultures taken | 8,175 |  | 16,217 |  | 120,023 |  | 106,419 |  | 12,913 |  | 34,588 |  | 12172 |  | 104,394 |  |
| No. of blood cultures per 1,000 patient days | 245.2 |  | 38.5 |  | 267.8 |  | 38.5 |  | 296.3 |  | 41.3 |  | 242.6 |  | 44.3 |  |
| Blood culture positivity rate (%) | 14.0 |  | 11.1 |  | 8.4 |  | 9.5 |  | 8.1 |  | 11.0 |  | 6.5 |  | 6.9 |  |
| No. of HOB episodes | 298 |  | 698 |  | 2,689 |  | 4,206 |  | 304 |  | 1,393 |  | 165 |  | 2,334 |  |
| No. of HOB episodes per 1,000 patient days | 8.9 |  | 1.7 |  | 6.0 |  | 1.5 |  | 7.0 |  | 1.7 |  | 3.2 |  | 1.0 |  |
| No. of HOB episodes with CC (% of all HOB) | 104 | (34.8) | 178 | (25.5) | 529 | (19.7) | 834 | (19.8) | 79 | (26.0) | 285 | (20.5) | 22 | (13.3) | 348 | (14.9) |
| No. of HOB episodes with CC per 1000 patient days | 3.1 |  | 0.4 |  | 1.2 |  | 0.3 |  | 1.8 |  | 0.3 |  | 0.4 |  | 0.2 |  |
| No. of HOB episodes with pathogen (% of all HOB) | 230 | (76.9) | 557 | (79.7) | 2,231 | (83.0) | 3,484 | (82.8) | 244 | (80.3) | 1,161 | (83.3) | 144 | (87.3) | 2,029 | (86.9) |
| No. of HOB episodes with pathogen per 1,000 patient days | 6.9 |  | 1.3 |  | 5.0 |  | 1.3 |  | 5.6 |  | 1.4 |  | 2.8 |  | 0.9 |  |
| No. of polymicrobial HOB episodes (% of all HOB) | 54 | (18.1) | 105 | (15.0) | 316 | (11.8) | 533 | (12.7) | 40 | (13.2) | 186 | (13.4) | 15 | (9.1) | 288 | (12.3) |
| No. of polymicrobial HOB episodes per 1,000 patient days | 1.6 |  | 0.2 |  | 0.7 |  | 0.2 |  | 0.9 |  | 0.2 |  | 0.3 |  | 0.1 |  |
| Median no. of in-hospital days until onset of HOB (IQR) | 12 | (7-22) | 12 | (6-23) | 19 | (9-38) | 12 | (5-23) | 12 | (6-21) | 9 | (4-17) | 21 | (7-22) | 29 | (7-33) |

CC = common commensal; HOB = hospital-onset bacteraemia and fungaemia; ICU = intensive care unit; IQR = interquartile range; No. = number.
Data depicted in this table pertains only to wards included in the study. Multiple HOB with different microorganisms occurring within a two-day period were counted as one polymicrobial HOB. Since polymicrobial HOB with both pathogen and common commensal were possible, the sum of HOB with pathogen and HOB with common commensal is greater than or equal to the number of all HOB. For counting blood cultures, only those taken from day two onwards were considered to align with HOB counting.

### Trends

**Figure 2A** shows HOB rates and the number of blood cultures per 1,000 patient days over time for all included in-patient areas separately for the four hospitals, while **Figure 2B** and **Figure 2C** pertain exclusively to data from ICU and non-ICU, respectively. In all hospitals, HOB rates seemed to align with blood culture frequency. When viewed over time, HOB rates remained rather stable at the respective hospitals. This was the case for all HOB combined, and for HOB with common commensals and HOB with pathogens considered separately. However, when looking at ICU only, HOB rates and blood culture frequency were higher during the years 2020 and 2021, coinciding with the COVID-19 pandemic.

**Figure 2.**
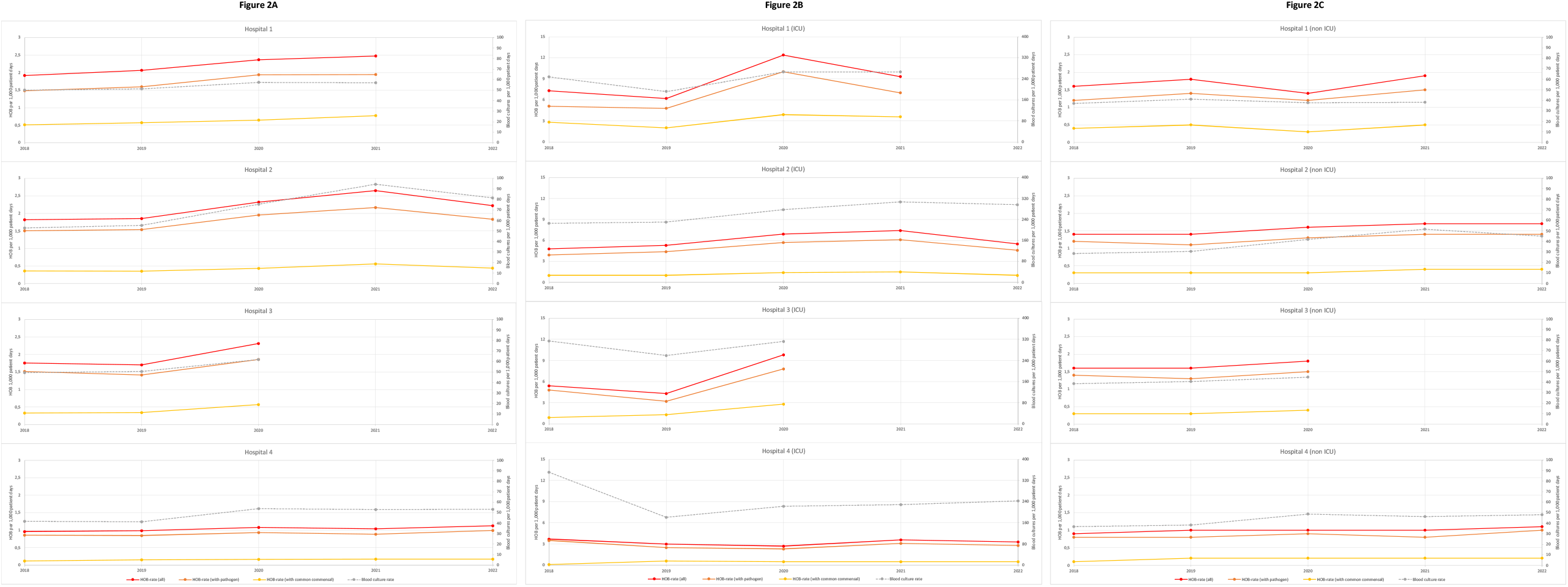
Rates of HOB and blood culture frequency over time per hospital HOB = hospital-onset bacteraemia and fungaemia; ICU = intensive care unit. Figure 2A pertains to all in-patient areas of the hospital. Figure 2B pertains only to intensive care units. Figure 2C pertains only to non-intensive care units.

### Causative microorganisms

Microorganisms in HOB showed variability between the four hospitals, with enterococci accounting for a larger portion of microorganisms in HOB in Hospital 1 than in the other hospitals and for example more Enterobacterales in Hospital 4. Moreover, differences between ICU and non-ICU were observed, with a general trend of more enterococci HOB in ICU across all four hospitals. Conversely, the proportion of HOB caused by Enterobacterales was higher in non-ICU than ICU. Of note, HOB caused by Staphylococcus aureus accounted for approximately 8.6% of HOB in Hospital 1, 9.1% in Hospital 2, 16.0% in Hospital 3, and 8.4% in Hospital 4. More details are depicted in **Figure 3**.

**Figure 3.**
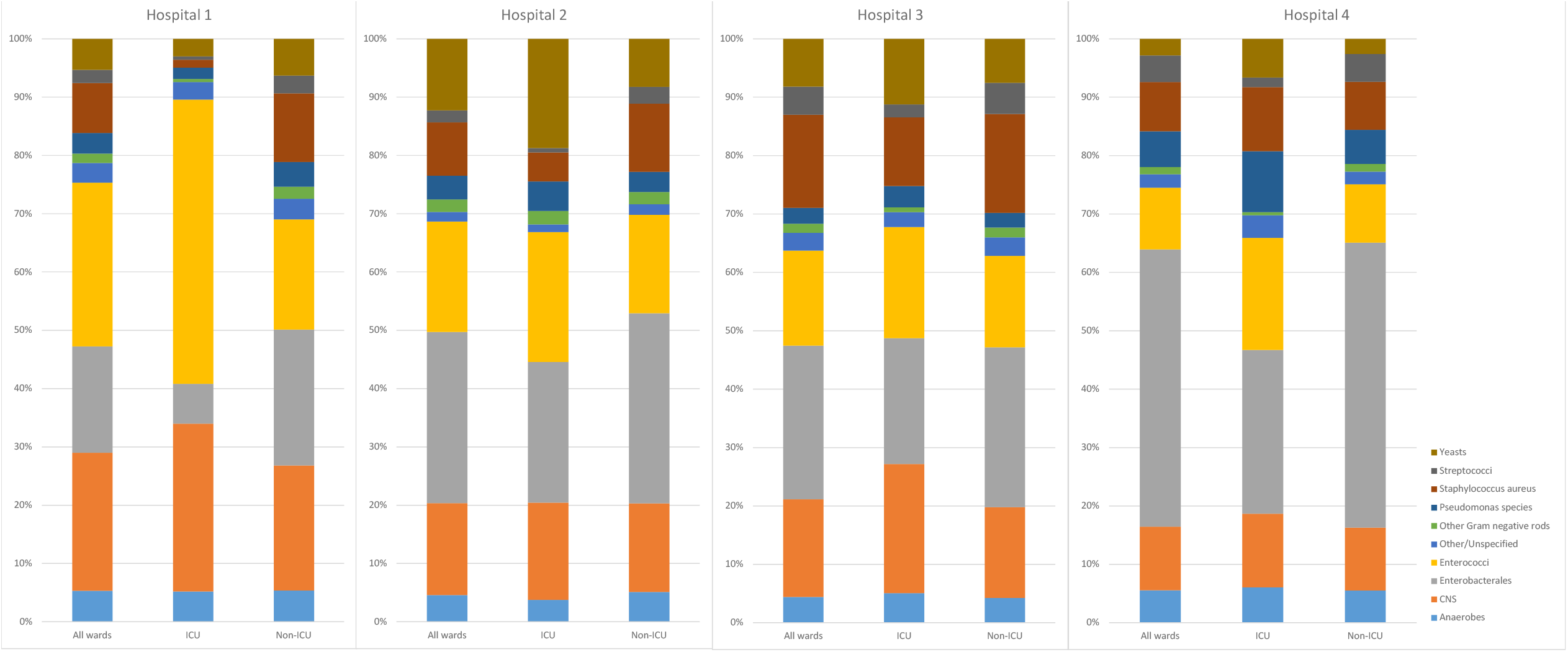
Share of microorganism group among all microorganisms in HOB episodes per hospital CNS = coagulase-negative staphylococci; ICU = intensive care unit

## Discussion

This multicentre retrospective cohort study is the first fully automated and standardised multinational implementation of HOB surveillance. We demonstrate the feasibility of developing and applying a consensus definition for the fully automated surveillance of HOB in four European hospitals and provide detailed guidance for future implementation. The incidence of HOB found in our study adds to results from other reports, ranging from 6.9 to 22.1 HOB per 1,000 patient days (3.2 to 8.9 in this study) in the adult ICU setting and around 1.8 per 1,000 patient days in the non-ICU setting (1.0 to 1.7 in this study) (10, 15). However, in many previous studies important details of the definition are not always provided or differ across studies, precluding direct comparison of rates and epidemiological characteristics (10, 13, 16, 17). In the PRAISE-HOB definition, we have addressed these possible sources of inconsistency by providing detailed implementation guidance.

The PRAISE-HOB consensus definition differs from recently published approaches in that single common commensals are never classified as HOB but discarded as solitary commensals, and the definition also specifies explicitly how to handle pre-existing community-onset bacteraemia. A further important element of the PRAISE-HOB definition that adds to previously published approaches, is the systematic grouping of microorganism episodes starting within 2 days of each other as single bacteraemia episodes (polymicrobial HOB), as this most likely represents a single clinical event. Interestingly, the contribution of polymicrobial HOB is sizeable, representing 12 to 16 % of HOB episodes in this study. Finally, we have set the episode duration (the timeframe during which onset of a new HOB with the same microorganisms is not possible) to 14 days, but other specifications (e.g., 30 days) can be found in the literature (18).

Application of the consensus definition to real-life data revealed both commonalities and differences in the epidemiology of HOB between four university hospitals in Europe. The patient population under consideration likely differed substantially across the four settings, hence the analysis does not aim to compare rates across hospitals. Strikingly however, the HOB rates were fairly consistent across the different hospitals and also the proportion of all bacteraemia episodes that were hospital-onset was comparable across the four hospitals. HOB surveillance may provide multiple insights to identify potential areas of quality improvement. For example, HOB surveillance allows for large-scale and consistent monitoring of the full breadth of the in-patient population. Interestingly, when looking at absolute numbers, the majority of HOB episodes in our study were attributable to non-ICU wards, likely resulting from the substantially higher number of non-ICU patient days. Nevertheless, the fact that HOB are not only occurring in ICU, and that in total numbers HOB are more frequent outside of ICU, is likely not exclusive to our study (19). This highlights the broad applicability of HOB surveillance to patients without central lines and of diverse underlying diseases, which represents a key advantage over CLABSI surveillance. HOB surveillance may also be employed to detect increased incidence of BSI over time. In our analysis, we observed an increase in HOB during the years most impacted by the COVID-19 pandemic (2020 and 2021), which is in alignment with other studies that have reported increased nosocomial BSI during the pandemic (20, 21). In addition, closer inspection of the local microorganism distribution can provide clues for targeted prevention efforts. For example, in Hospitals 1 and 4 respectively, the relatively large share of enterococci or Gram-negative HOB may lead to further investigations and pathogen-specific HOB rates (e.g. S. aureus or Candida sp.) may also provide information that can inform IPC responses.

In this study we did not perform an assessment of the sources of HOB. Recent studies have found that major sources include endovascular infections, gastro-intestinal and abdominal infections, urinary tract infections, skin and soft tissue or surgical site infections, while in a considerable proportion, no known source could be detected (16). Interestingly, although there seems to be correlation between HOB and CLABSI rates, their overlap appears to be only modest. In a recent study from India, only 13% of HOB represented CLABSI episodes (15). Conversely, HOB rates showed a high correlation with CLABSI rates in a study focused on the American ICU setting (10). To gain deeper insights into both source of HOB and its concordance with established surveillance metrics such as CLABSI, further research will be necessary, particularly with a focus on the epidemiology in Europe. Another aspect warranting further study is the preventability of HOB. Several studies have described and used methods to assess the preventability of HOB (22) and initial results demonstrate that over 50% of HOB might be potentially preventable (15, 16, 23). Application of uniform definitions and methods across hospitals and countries, as demonstrated in this study, will likely be of great value when assessing preventability and characterising the clinical significance of HOB episodes. Similarly, future research is needed to determine the most suitable approach for reporting HOB-related results to stimulate improvements in the quality of care, and suitable means of benchmarking HOB data, taking into account extraneous factors affecting HOB rates, such as blood culture frequency or baseline risk.

The detailed consensus definition and detailed implementation guidance provided in this study allows for implementation in settings with different IT infrastructure. We expect it to also be feasible outside of the university hospital setting, as the data needed for implementation are likely available in most European EHR systems. Importantly however, giving priority to developing fully automated surveillance definition that is feasible in variable settings, meant that we purposefully excluded clinical details, possibly at the cost of accuracy in some cases. Given the intended broad applicability of the PRAISE HOB consensus definition, we believe it will serve as an important reference in the context of transitioning from manual to automated surveillance in Europe, and that it will be complementary to other activities on the matter at the national and international level in Europe (24, 25).

This study has several limitations. The specifications of the HOB definitions were defined based on consensus discussion, supported by existing literature and sometimes by exploratory data analysis. The impact and suitability of certain choices (e.g., duration of episode, definition for polymicrobial HOB) on the outcomes, will have to be investigated in follow-up studies. In addition, the retrospective data analysis was limited to existing patient cohorts, thereby leading to not fully overlapping time periods between the four hospitals. Finally, at this stage of development, we did not include information on antimicrobial susceptibility into the algorithm and the data presented excludes the paediatric population. Moreover, as manual HOB surveillance was not performed in our centres using a similar definition, we did not validate the automated HOB algorithm against manual surveillance and we also did not validate our results directly against data show in EHR systems.

In conclusion, the definition and method for automated HOB surveillance was successfully applied and reproduceable in four hospitals from four different European countries. This new consensus definition addresses the difficulty to transpose the existing definitions for surveillance to automated surveillance and provides an important basis for future development. Future studies will need to further characterise the conditions underlying HOB episodes, assess preventability and effective interventions, and define the metric’s role in infection prevention programs.

## Declarations

### Ethical statement

In all hospitals ethical approval was obtained separately and informed consent was not needed; if applicable per local regulations patients who objected to the use of their data for the purpose of research were excluded.

Process numbers of ethical approval are as follows: UMC Utrecht: 21-856DB; Charité-Universitätsmedizin Berlin: EA2/060/23; Region Stockholm: 2018/1030-31.

The data collection and analysis of the Swiss cohort at the Hôpitaux Universitaires de Genève (HUG) was considered as falling outside of the scope of the Swiss legislation regulating research on human subjects, so that the need for local ethics committee approval was waived by the Commission Cantonale d^′^Éthique de la Rercherche de Genève (decision number: Req-2024-01048).

## Use of artificial intelligence tools

None declared.

## Data availability

The datasets generated and/or analysed during the current study are not publicly available due to ethical limitations related to sharing patient information, but are available via the corresponding author on reasonable request.

## Acknowledgements

Dr Aghdassi is participant in the Charité Digital Clinician Scientist Program funded by the DFG, the Charité Universitätsmedizin – Berlin, and the Berlin Institute of Health at Charité (BIH).

## Authors’ contributions

All authors and collaborators were involved in drafting the consensus definition and minimal dataset. MB, LP, DS, SvdW, and DT performed data collection and applied the algorithm. All authors were involved in data interpretation. MvM, SA, SvdW drafted the initial manuscript. All authors and collaborators reviewed and approved the manuscript.

