## Appendix 1 for "Hospital-onset bacteraemia and fungaemia as a novel automated surveillance indicator: results from four European university hospitals"

#### PRAISE-HOB Consensus definition & Minimal dataset

**This supplement describes the definition of hospital-onset bacteraemia and fungaemia (HOB) as defined by the PRAISE (Providing a Roadmap for Automated Infection Surveillance in Europe) network.**

**Authors**

Seven J.S. Aghdassi [1,2*], Suzanne D. van der Werff [3,4*], Gaud Catho [5,6], Manon Brekelmans [7,8], Luis A. Peña Diaz [1], Niccolò Buetti [5,9], Ferenc D. Rüther [1], Daniel Dinis Teixeira [5], Daniel Sjöholm [10], Pontus Nauclér [3,4], Michael Behnke [1#], Maaike S.M. van Mourik [7#], on behalf of the PRAISE-HOB working group

*;# These authors contributed equally to this work and share first/last authorship.

**Affiliations**

1. Charité – Universitätsmedizin Berlin, corporate member of Freie Universität Berlin and Humboldt-Universität zu Berlin, Institute of Hygiene and Environmental Medicine, Berlin, Germany
2. Berlin Institute of Health at Charité – Universitätsmedizin Berlin, BIH Biomedical Innovation Academy, BIH Charité Digital Clinician Scientist Program, Berlin, Germany
3. Department of Medicine Solna, Division of Infectious Diseases, Karolinska Institutet, Stockholm, Sweden
4. Department of Infectious Diseases, Karolinska University Hospital, Stockholm, Sweden
5. Infection Control Program and WHO Collaborating Centre, University of Geneva Hospitals and Faculty of Medicine, Geneva, Switzerland
6. Infectious diseases division, Central Institute, Valais Hospital, Sion, Switzerland
7. Department of Medical Microbiology and Infection Control, University Medical Centre Utrecht, the Netherlands
8. Centre for Infectious Diseases Control, National Institute for Public Health and the Environment, Bilthoven, the Netherlands
9. Université Paris Cité, Inserm, IAME, Paris, France
10. Department of Medicine Solna, Division of Clinical Epidemiology, Karolinska Institutet, Stockholm, Sweden

**Version: 1.0**

**Date: 2024-07-19**

Always acquire latest version via PRAISE Network

### Table of contents

PRAISE-HOB Consensus definition 3
Flow chart of the PRAISE-HOB algorithm 5
Examples of the PRAISE-HOB algorithm 6
Minimal dataset (input) of the PRAISE-HOB algorithm 13
Stage 1 13
Stage 2 14
Look-up tables 15
Microorganism classification (based on CDC Common commensals). 16
Ward classification 16
Minimal dataset (output) of the PRAISE-HOB algorithm 17
Examples for denominator calculation 19
Example of the midnight method for use of patient days as denominator 19
Examples of denominator data analysis by wardGroupType and wardGroupValue 19

Always acquire latest version via PRAISE Network

### PRAISE-HOB Consensus definition

The PRAISE-HOB definition classifies blood cultures of in inpatients according to standardized definitions.

*Key concepts in the PRAISE-HOB algorithm and definition*

| | |
|---|---|
| **Hospital admission** | A hospital admission is defined as at least one overnight stay. The day of admission is considered day 0. An admission ends upon discharge. |
| **Blood culture** | Blood cultures are counted at the blood culture 'set' level (sample ID). Hence, 1 blood culture may consist of 1 or 2 vials. Sample date is considered day 0.<br>Blood cultures are eligible if collected during hospital admission[1.] If a patient is readmitted on the same day, all blood cultures from that day are related to the prior admission. |
| **Microorganism in blood culture** | All microorganisms (bacteria and or fungi) isolated from eligible blood cultures. |
| **Common commensals and pathogens** | Common commensals are defined in line with the current CDC/NHSN classification[2]. All microorganisms not classified as common commensal are considered pathogens. |
| **Microorganism event** | Defined by either a pathogen in 1 blood culture OR the same common commensal (genus + species) in 2 blood cultures (different sample ID's) within 2 calendar days of each other (i.e., day 0, 1, 2). Results of antimicrobial susceptibility testing are not included in determining similarity.<br>Date of microorganism event = sampling date of the 1st culture with pathogen or 1st culture with a common commensal that is part of a set of at least 2. |
| **Microorganism episode** | For each microorganism event, a subsequent 14-day period is defined, lasting from day 0 – 13 relative to the microorganism event OR until hospital discharge.<br>The start date of a microorganism episode is the date of the microorganism event. The end date of the microorganism episode is 13 days after the microorganism event or at hospital discharge, whichever comes first. |
| **Copy strains** | All microorganisms of the same species isolated from blood cultures of the same patient and taken within the microorganism episode timeframe (and hence the same hospital admission) are classified as copy strains. |
| **Solitary commensal** | A common commensal that is not confirmed by a second blood culture within 2 calendar days (i.e. day 0, 1, or 2) AND is not a copy strain is classified as a solitary commensal. |

Always acquire latest version via PRAISE Network

| | |
|---|---|
| **Bacteraemia episode** | One or several microorganism episodes can constitute a bacteraemia episode. Microorganism episodes with start dates within 2 calendar days (i.e. day 0, 1, or 2) of each other are grouped together as one bacteraemia episode. The start date of the bacteraemia episode is the date of the first microorganism episode.<br><br>If there is only a single microorganism episode, this is a **monomicrobial** bacteraemia episode. In that case, the start and end date of the bacteraemia episode coincide with the dates of the microorganism episode.<br><br>If there are 2 or more microorganism episodes within 2 calendar days (i.e., day 0, 1, or 2) of each other, these are **polymicrobial** bacteraemia episodes. Here, the start date of the bacteraemia episode is the start date of the first microorganism episode included, and the end date of the bacteraemia episode is the end date of the last included microorganism episode. Consequently, the maximum possible duration of a bacteraemia episode is 16 days.<br>Of note, this does not influence copy strain elimination which happens at the microorganism episode level. |
| **Hospital-onset bacteraemia** | A bacteraemia episode qualifies as HOB if the start date of the bacteraemia episode is >=2 days after admission. Admissions start on day 0, so any bacteraemia episode starting on day 2 of admission or later is classified as a HOB. |
| **Community-onset bacteraemia** | Bacteraemia episodes starting on day 0 or 1 of admission are classified as community-onset bacteraemia. |
| **Attributable ward** | The bacteraemia episode is attributed to the ward where the patients was 2 calendar days prior to the start date of the bacteraemia episode. If the patient was transferred on that day, the ward where the patient was first is selected. |
| **Ward specialty** | Ward specialties are classified according to the ECDC ward specialty classification. Patient specialty classification is optional (see minimal dataset). |
| **Patient days** | For the denominator, patient days are calculated by ward specialty. This employs the midnight method, meaning that the ward where the patient was at midnight (00:00) is counted in the denominator (see example in MDS).<br><br>The entire admission is included, also the first and last two days of admission. |

[1] Blood cultures taken in the emergency room are included within the hospital admission for the purpose of applying the algorithm.

[2] Common commensal classification used in this version is from 2023. We recommend using the most recent version when implementing algorithms.

Notes:

- Time intervals are counted in calendar days for reasons of feasibility.
- Some patients may have positive blood cultures in outpatient settings prior to admission, be transferred from other facilities with a prolonged bacteraemia, or be readmitted soon after discharge with positive blood cultures. These are relatively infrequent scenarios, and we did not include these for the sake of clarity of interpretation.
- In all place where bacteraemia is mentioned, this also includes fungaemia.

Always acquire latest version via PRAISE Network

### Flow chart of the PRAISE-HOB algorithm

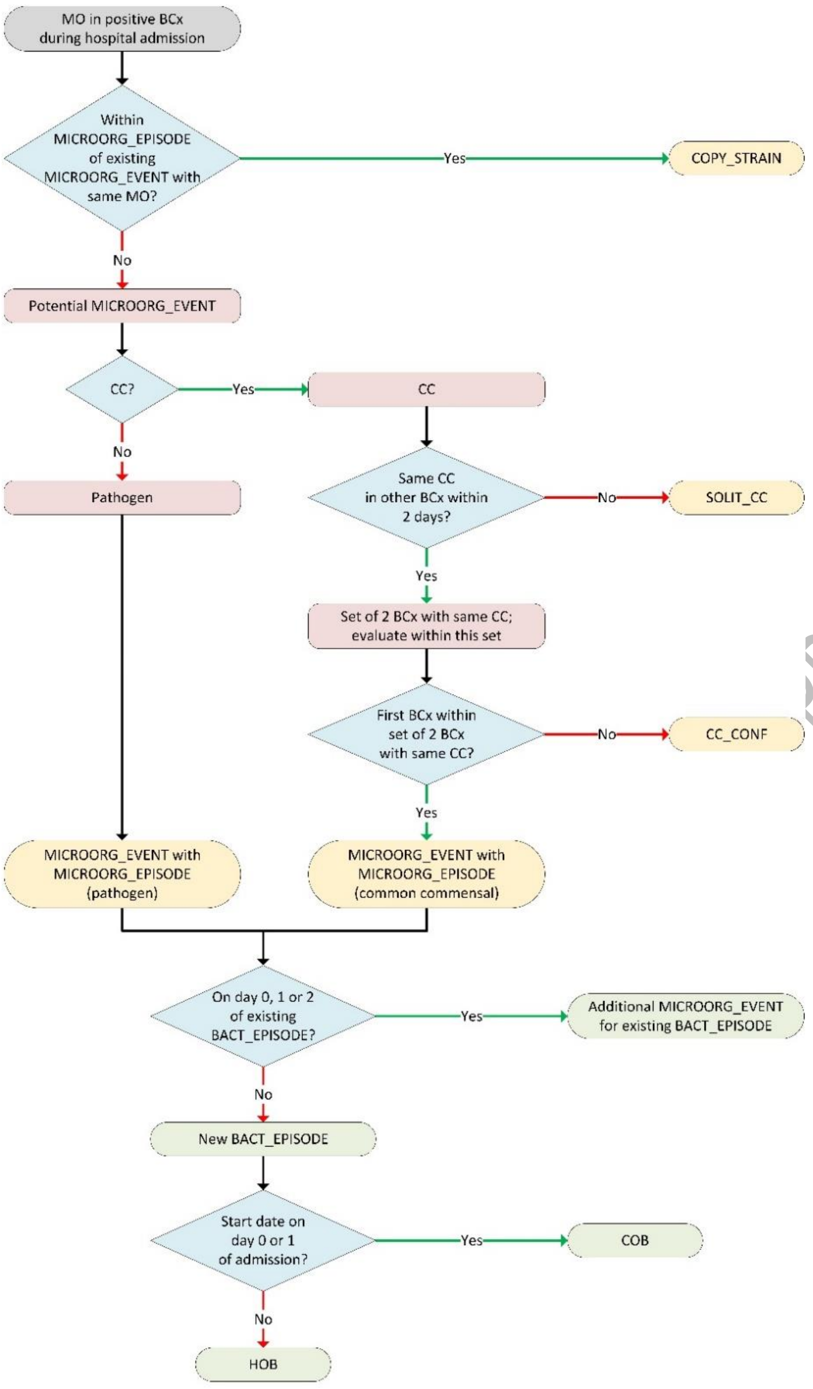

PRAISE Network

**Abbreviations/explanations**

**MO**: microorganism, only bacteria and fungi, on genus and species level
**BCx**: blood culture (on cultureID level)
**CC**: common commensal, based on CDC/NHSN list
**MICROORG_EVENT**: microorganism event; pathogen in BCx or common commensal in BCx confirmed by second BCx with same common commensal within 2 calendar days (day 0, 1 & 2)
**CC_CONF**: common commensal confirmation BCx; second BCx with same common commensal within 2 days (day 0-2) of previous BCx to confirm MICROORG_EVENT for common commensal
**COPY_STRAIN**: repeated MO in BCx within MICROORG_EPISODE period of MICROORG_EVENT
**MICROORG_EPISODE**: microorganism episode; MICROORG_EVENT (first BCx), CC_CONF (second BCx for common commensal) and COPY STRAINs within 14-day period (day 0-13) of start MICROORG_EVENT
**SOLIT_CC**: solitary common commensal; common commensal in BCx without CC_CONF and not within MICROORG_EPISODE of MICROORG_EVENT
**BACT_EPISODE**: bacteraemia and fungaemia episode; MICROORG_EVENTS with their associated MICROORG_EPISODEs which occurred within 2 days of each other (day 0, 1 & 2)
**COB**: community-onset bacteraemia and fungaemia; BACT_EPISODE started on day 0 or 1 of admission
**HOB**: hospital-onset bacteraemia and fungaemia; BACT_EPISODE started on day 2 or later of admission

Notes:
This flowchart assumes that microorganism identified in blood cultures are processed independently and sorted in chronological order. This flowchart is not meant for real-time processing.

#### Examples of the PRAISE-HOB algorithm

The following examples illustrate the PRAISE-HOB algorithm in different scenarios. All examples are based on fictitious data.

1. HOB with a single microorganism

| Hospital admission day | 0 | 1 | 2 | 3 | 4 | 5 | 6 | 7 | 8 | 9 | 10 | 11 | 12 |
|---|---|---|---|---|---|---|---|---|---|---|---|---|---|
| Microorg. from BC | | | | *E. coli* | | | | | | | | | |
| Ward name | Ward 1 | Ward 1 | Ward 1 | Ward 1 | Ward 1 | Ward 1 | Ward 1 | Ward 1 *Hospital discharge at 15:00* | | | | | |
| Ward specialty | MED | MED | MED | MED | MED | MED | MED | MED | x | x | x | x | x |

Conclusion of algorithm: 1 HOB with *E. coli* attributable to Ward 1 of ward specialty MED. The HOB-episode starts on Day 3 and ends with hospital discharge on Day 7.

2. HOB with common commensal

| Hospital admission day | 0 | 1 | 2 | 3 | 4 | 5 | 6 | 7 | 8 | 9 | 10 | 11 | 12 |
|---|---|---|---|---|---|---|---|---|---|---|---|---|---|
| Microorg. from BC | *S. hominis* | | | | | *S. epidermidis in 2 BC sets* | | | | | | | |
| Ward name | Ward 1 | Ward 1 | Ward 1 | Ward 1 | Ward 1 | Ward 1 | Ward 1 | Ward 1 *Hospital discharge at 15:00* | | | | | |
| Ward specialty | MED | MED | MED | MED | MED | MED | MED | MED | x | x | x | x | x |

Conclusion of algorithm: 1 HOB with *S. epidermidis* attributable to Ward 1 of ward specialty MED. The HOB-episode starts on Day 5 and ends with hospital discharge on Day 7.

*Note: Both S. epidermidis and S. hominis are common commensals. Blood culture with S. hominis (on day 0, admission day) is considered a solitary commensal.*

Always acquire latest version via PRAISE Network

3. HOB with pathogen AND solitary commensal

| Hospital admission day | 0 | 1 | 2 | 3 | 4 | 5 | 6 | 7 | 8 | 9 | 10 | 11 | … | 17 | 18 |
|---|---|---|---|---|---|---|---|---|---|---|---|---|---|---|---|
| Microorg. from BC | | | | | *S. aureus*<br>*S. epidermidis* | | | | | | | | | | |
| Ward name | Ward 1 | Ward 1 | Ward 1 *Transfer to Ward 2 at 3 pm* | Ward 2 | Ward 2 | Ward 2 *Transfer to ward 1 at 16:00* | Ward 1 | Ward 1 | Ward 1 | Ward 1 | Ward 1 | Ward 1 | … | Ward 1 | Ward 1 *Hospital discharge at 15:00* |
| Ward specialty | SUR | SUR | SUR | ICU | ICU | ICU | SUR | SUR | SUR | SUR | SUR | SUR | … | SUR | SUR |

Conclusion of algorithm: 1 HOB with *S. aureus* attributable to Ward 1 of ward specialty SUR. The HOB-episode starts on Day 4 and ends on Day 17.
*Note: The attribution to Ward 1 is due to the fact that, two days before the start of the HOB-episode, the patient was first present on Ward 1 and then on Ward 2. S. epidermidis is considered a solitary commensal.*

4. Polymicrobial HOB with 2 pathogens, 2 days apart

| Hospital admission day | 0 | 1 | 2 | 3 | 4 | 5 | 6 | 7 | 8 | 9 | 10 | 11 | 12 | … | 26 | 27 |
|---|---|---|---|---|---|---|---|---|---|---|---|---|---|---|---|---|
| Microorg. from BC | | | | | | | *E. coli* | | *S. marcescens* | | | | | | | |
| Ward name | Ward 1 | Ward 1 | Ward 1 | Ward 1 | Ward 1 | Ward 1 *Transfer to Ward 2 at 11:00* | Ward 2 | Ward 2 | Ward 2 | Ward 2 | Ward 2 *Transfer to Ward 1 at 12:00* | Ward 1 | Ward 1 | … | Ward 1 | Ward 1 *Hospital discharge at 15:00* |
| Ward specialty | SUR | SUR | SUR | SUR | SUR | SUR | MIX | MIX | MIX | MIX | MIX | SUR | SUR | … | SUR | SUR |

Conclusion of the algorithm: 1 polymicrobial HOB with *E. coli* and *S. marcescens* attributable to Ward 1 of ward specialty SUR. The HOB-episode starts on Day 6 and ends on Day 21.
*Note: Since S. marcescens is part of a polymicrobial HOB-episode starting on Day 6, the whole episode is attributed to Ward 1 = SUR (-2 days relative to Day 6). The polymicrobial bacteraemia episode starts on day 6 with the microorganism episode of E. coli and terminates after the end of the microorganism episode of S. marcescens, which is on Day 21. Consequently, a new HOB with an E. coli microorganism episode on Day 20 or Day 21 would be possible despite still being within the timeframe of the HOB-episode. This is due to the fact that copy strain elimination happens on the microorganism episode level (see definition table).*

Always acquire latest version via PRAISE Network

5. Community-onset bacteraemia on admission, and cultures with the same pathogen identified later during admission.

| Hospital admission day | 0 | 1 | 2 | 3 | 4 | 5 | 6 | 7 | 8 | 9 | 10 | 11 | 12 |
|---|---|---|---|---|---|---|---|---|---|---|---|---|---|
| Microorg. from BC | *S. aureus* | | | | *S. aureus* | | | | | | | | |
| Ward name | Ward 1 | Ward 1 | Ward 1 | Ward 1 | Ward 1 | Ward 1 | Ward 1 | Ward 1 | Ward 1 | Ward 1 | Ward 1 | Ward 1 | Ward 1 *Hospital discharge at 15:00* |
| Ward specialty | MED | MED | MED | MED | MED | MED | MED | MED | MED | MED | MED | MED | MED |

Conclusion of algorithm: No HOB. *Note: This patient has a community-onset bacteraemia episode that started on Day 0, and hence is not hospital-onset. S. aureus on Day 4 is a copy strain in the bacteraemia episode starting on admission. The community-onset microorganism episode ends with hospital discharge on Day 12.*

6. Pre-existing bacteraemia on admission and new cultures with a different pathogen

| Hospital admission day | 0 | 1 | 2 | 3 | 4 | 5 | 6 | 7 | 8 | 9 | 10 |
|---|---|---|---|---|---|---|---|---|---|---|---|
| Microorg. from BC | *S. aureus* | | | | | | *E. coli* | | | | |
| Ward name | Ward 1 | Ward 1 | Ward 1 *Transfer to ward 2 at 10:00* | Ward 2 | Ward 2 | Ward 2 | Ward 2 | Ward 2 | Ward 2 | Ward 2 | Ward 2 *Hospital discharge at 15:00* |
| Ward specialty | ICU | ICU | ICU | MED | MED | MED | MED | MED | MED | MED | MED |

Conclusion of algorithm: 1 HOB with *E. coli* attributable to Ward 2 of ward specialty MED. The HOB-episode starts on Day 6 and ends with the hospital discharge on Day 10.
*Note: Additionally, this patient has a community-onset bacteraemia episode with S. aureus that started on Day 0 and ends with hospital discharge on Day 10.*

Always acquire latest version via PRAISE Network

7. Prolonged bacteraemia

| Hospital admission day | 0 | 1 | 2 | 3 | 4 | 5 | 6 | 7 | 8 | 9 | 10 | 11 | 12 | 13 | 14 | 15 | 16 | 17 | … | 40 |
|---|---|---|---|---|---|---|---|---|---|---|---|---|---|---|---|---|---|---|---|---|
| Microorg. from BC | *S. aureus* | | | | | | | | *S. aureus* | | | | | | | | *S. aureus* | | | |
| Ward name | A | A | A | A | A | A | A | A | A | A | A | A | A | A | A | A | A | A | A | A<br>*Hospital discharge at 15:00* |
| Ward specialty | MED | MED | MED | MED | MED | MED | MED | MED | MED | MED | MED | MED | MED | MED | MED | MED | MED | MED | MED | MED |

Conclusion of algorithm: 1 HOB with *S. aureus* attributable to Ward A of ward specialty MED. The HOB-episode starts on Day 16 and ends on Day 29.
*Note: S. aureus on Day 8 is a copy strain within the microorganism episode on admission (community-onset bacteraemia episode). S. aureus on Day 8 does not extent the microorganism episode. Therefore, S. aureus on Day 16 occurs outside of the 14-day microorganism episode window and marks the start of a new microorganism episode.*

8. Prolonged bacteraemia after a polymicrobial HOB (extreme example)

| Hospital admission day | 0 | 1 | 2 | 3 | 4 | 5 | 6 | 7 | 8 | 9 | 10 | 11 | 12 | 13 | 14 | 15 | 16 | 17 | 18 | 19 |
|---|---|---|---|---|---|---|---|---|---|---|---|---|---|---|---|---|---|---|---|---|
| Microorg. from BC | *S. aureus* | | *E. coli* | | | | | | | | | | | | *E. coli*<br>*S. aureus* | | | | | |
| Ward name | A | A | A | A | A | A | A | A | A | A | A | A | A | A | A | A | A | A | A | A<br>*Hospital discharge at 15:00* |
| Ward specialty | MED | MED | MED | MED | MED | MED | MED | MED | MED | MED | MED | MED | MED | MED | MED | MED | MED | MED | MED | MED |

Conclusion of algorithm: 1 HOB with *S. aureus* attributable to Ward A of ward specialty MED. The HOB-episode starts on Day 14 and ends with hospital discharge on Day 19.
*Note: This patient has a polymicrobial community-onset bacteraemia episode with S. aureus and E. coli. The E. coli on Day 14 falls within the E. coli microorganism episode starting on Day 2. The E. coli on Day 14 is hence a copy strain. The S. aureus on Day 14 falls outside of the of S. aureus microorganism episode starting on Day 0, hence constitutes a HOB (and not a copy strain).*

Always acquire latest version via PRAISE Network

9. Polymicrobial HOB with two common commensals

| Hospital admission day | 0 | 1 | 2 | 3 | 4 | 5 | 6 | 7 | 8 | 9 | 10 |
|---|---|---|---|---|---|---|---|---|---|---|---|
| Microorg. from BC | | | | *S. epidermidis* | *S. hominis*<br>*S. epidermidis* | | *S. hominis* | | | | |
| Ward name | Ward 1 | Ward 1 | Ward 1 | Ward 1 | Ward 1<br>*Transfer to Ward 2 at 14:00* | Ward 2 | Ward 2 | Ward 2 | Ward 2 | Ward 2 | Ward 2<br>*Hospital discharge at 15:00* |
| Ward specialty | ICU | ICU | ICU | ICU | ICU | SUR | SUR | SUR | SUR | SUR | SUR |

Conclusion of algorithm: 1 polymicrobial HOB with *S. epidermidis and S. hominis* attributable to Ward 1 of ward specialty ICU. The HOB-episode starts on Day 3 and ends with hospital discharge on Day 10.

*Note: Both S. epidermidis and S. hominis are common commensals. The two microorganism episodes are merged into a polymicrobial HOB since they start within 2 days of each other.*

10. Overlapping HOB-episodes

| Hospital admission day | 0 | 1 | 2 | 3 | 4 | 5 | 6 | 7 | 8 | … | 25 |
|---|---|---|---|---|---|---|---|---|---|---|---|
| Microorg. from BC | | | | *S. aureus* | | | *E. coli* | | | | |
| Ward name | Ward 1 | Ward 1 | Ward 1 | Ward 1 | Ward 1<br>*Transfer to Ward 2 at 10:00* | Ward 2 | Ward 2 | Ward 2 | Ward 2 | … | Ward 2<br>*Hospital discharge at 15:00* |
| Ward specialty | ICU | ICU | ICU | ICU | ICU | MED | MED | MED | MED | … | MED |

Conclusion of algorithm: 2 HOB

1 HOB with *S. aureus* attributable to Ward 1 of ward specialty ICU. This HOB-episode starts on Day 3 and ends on Day 16.

1 HOB with *E. coli* attributable to Ward 1 of ward specialty ICU. This HOB-episode starts on Day 6 and ends on Day 19.

*Note: The two microorganism episodes are not merged into a polymicrobial HOB because they do not start within 2 days of each other.*

Always acquire latest version via PRAISE Network

11. Polymicrobial HOB and overlapping HOB-episode.

| Hospital admission day | 0 | 1 | 2 | 3 | 4 | 5 | 6 | 7 | 8 | 9 | 10 | … | 30 |
|---|---|---|---|---|---|---|---|---|---|---|---|---|---|
| Microorg. from BC | | | | *E. coli*<br>*S. epidermidis*<br>*S. epidermidis* | *S. aureus* | *S. hominis* | | | | *S. hominis* | *S. hominis* | | |
| Ward name | Ward 1 | Ward 1 | Ward 1 | Ward 1 | Ward 1 | Ward 1 | Ward 1 | Ward 1 | Ward 1 | Ward 1 | Ward 1 | … | Ward 1<br>*Hospital discharge at 15:00* |
| Ward specialty | MED | MED | MED | MED | MED | MED | MED | MED | MED | MED | MED | … | MED |

Conclusion of algorithm: 2 HOB

1 polymicrobial HOB with *E. coli*, *S. epidermidis* and *S. aureus* attributable to Ward 1 of ward specialty MED. This HOB-episode starts on Day 3 and ends on Day 17.

1 HOB with *S. hominis* attributable to Ward 1 of ward specialty MED. This HOB-episode starts on Day 9 and ends on Day 22.

*Note: S. epidermidis and S. hominis are common commensals. The S. hominis on Day 5 is a solitary commensal. The polymicrobial bacteraemia episode starts on day 3 with the microorganism episodes of E. coli & S. epidermidis and terminates after the end of the microorganism episode of S. aureus, which is on Day 17. Consequently, a new HOB with an E. coli and/or S. epidermidis microorganism episode on Day 17 would be possible despite still being within the timeframe of the HOB-episode. This is due to the fact that copy strain elimination happens on the microorganism episode level (see definition table).*

12. Solitary commensals do not make a HOB.

| Hospital admission day | 0 | 1 | 2 | 3 | 4 | 5 | 6 | 7 | 8 | 9 | 10 |
|---|---|---|---|---|---|---|---|---|---|---|---|
| Microorg. from BC | | | | | | | *S. epidermidis*<br>*S. hominis* | *Micrococcus luteus* | | | |
| Ward name | Ward 1 | Ward 1 | Ward 1 | Ward 1 | Ward 1<br>*Transfer to Ward 2 at 10:00* | Ward 2 | Ward 2 | Ward 2 | Ward 2 | Ward 2 | Ward 2<br>*Hospital discharge at 15:00* |
| Ward specialty | ICU | ICU | ICU | ICU | ICU | MED | MED | MED | MED | MED | MED |

Conclusion of algorithm: 0 HOB. *Note: All identified microorganisms are common commensals. Different solitary commensals do not constitute a HOB. All identified microorganisms are considered solitary commensals.*

Always acquire latest version via PRAISE Network

13. “A chain of positive BC” (extreme example)

| Hospital admission day | 0 | 1 | 2 | 3 | 4 | 5 | 6 | 7 | 8 | 9 | 10 | … | 30 |
|---|---|---|---|---|---|---|---|---|---|---|---|---|---|
| Microorg. from BC | | | | *S. aureus* | *E. coli* | | *P. aeruginosa* | *E. coli* | | *K. pneumoniae* | | | |
| Ward name | Ward A | Ward A | Ward A | Ward A | Ward A | Ward A | Ward A | Ward A | Ward A | Ward A | Ward A | … | Ward A<br>*Hospital discharge at 15:00* |
| Ward specialty | MED | MED | MED | MED | MED | MED | MED | MED | MED | MED | MED | … | MED |

Conclusion of algorithm: 3 HOB.

1 polymicrobial HOB with *S. aureus* and *E. coli* attributable to Ward A of ward specialty MED. This HOB-episode starts on Day 3 and ends on Day 17.

1 HOB with *P. aeruginosa* attributable to Ward A of ward specialty MED. This HOB-episode starts on Day 6 and ends on Day 19.

1 HOB with *K. pneumoniae* attributable to Ward A of ward specialty MED. This HOB-episode starts on Day 9 and ends on Day 22.

*Note: The polymicrobial bacteraemia episode starts on day 3 with the microorganism episodes of S. aureus and terminates after the end of the microorganism episode of E. coli, which is on Day 17. Consequently, a new HOB with an S. aureus microorganism episode on Day 17 would be possible despite still being within the timeframe of the HOB-episode. This is due to the fact that copy strain elimination happens on the microorganism episode level (see definition table).*

*P. aeruginosa is more than two days after the first microorganism episode of the polymicrobial HOB and therefore not added to it. E. coli on Day 7 is a copy strain and is therefore not merged with P. aeruginosa on Day 6 to form a polymicrobial HOB. K. pneumonia and P. aeruginosa are more than two days apart and therefore not merged into a polymicrobial episode.*

14. No post-discharge surveillance

| Hospital admission day | Jan 1 | Jan 2 | Jan 3 | Jan 4 | Jan 5 | … | Jan 12 | Jan 13 | Jan 14 | Jan 15 | … | Jan 29 |
|---|---|---|---|---|---|---|---|---|---|---|---|---|
| Microorg. from BC | | | | *S. aureus* | *S. epidermidis in 2 BC sets* | | | | *S. aureus* | | | |
| Ward name | Ward 1 | Ward 1 | Ward 1 | Ward 1<br>*Transfer to other facility* | | | Ward 1<br>(*Hospital readmission)*<br>*Then transfer to ICU at 17:00* | Ward 2 | Ward 2<br>*Transfer to Ward 1 at 16:00* | Ward 1 | … | Ward 1<br>*Hospital discharge at 15:00* |
| Ward specialty | MED | MED | MED | MED | | | MED | ICU | ICU | MED | … | MED |

Conclusion of algorithm: 2 HOB.

1 HOB with *S. aureus* during the first hospital stay attributable to Ward 1 of ward specialty MED. This HOB-episode starts on January 4 and ends with the hospital discharge (transfer) on January 4.

1 HOB with *S. aureus* during the second hospital stay attributable to Ward 1 of ward specialty MED. This HOB-episode starts on January 14 and ends on January 27.

*Note: The 2 S. epidermidis on January 5 are not considered because it was taken outside of an in-patient admission. S. aureus on January 14 is not a copy strain of the S. aureus HOB-episode of January 4 since these are separate hospital admissions and HOB-episodes are not “carried over” between separate admissions.*

Always acquire latest version via PRAISE Network

#### Minimal dataset (input) of the PRAISE-HOB algorithm

In the tables below we describe the minimal dataset agreed upon. This dataset describes the true minimum input to achieve the output file and allow for reporting as shown in the manuscript. An excel file describing the minimal dataset is also available.

**Stage 1**: this describes the minimal dataset for hospitals/data providers that cannot locally link data needed by the algorithm (e.g., linking of blood cultures to wards).

**Stage 2:** for hospitals/data providers able to perform linkage of data files: they can perform linkage locally and only provide the data needed by the algorithms.

Note: it is not envisioned to mix stage 1 and 2. It is advised to either provide stage 1 or stage 2 data.

##### Stage 1

| Variable | Data type | Values or format (if applicable) | Optional? | Comments |
|---|---|---|---|---|
| **PATIENTS.CSV** | | | | For each patient with HOB |
| patientId | string | | MANDATORY | Should be pseudonymised |
| patientSex | string | M (male); F (female); X (not male not female); U (unknown) | MANDATORY | |
| patientBirthDate | date | YYYY/MM/DD | MANDATORY | For sharing purpose consider dummy |
| patientDeathDate | date | YYYY/MM/DD | Optional | |
| **BLOODCULTURES.CSV** | **(1 line per microorganism)** | | | *Includes positive AND negative blood cultures.* |
| bcId | string | | MANDATORY | A unique identifier of BC microorganism. For example compound sampleId+isolateNumber. Must add isolateNumberfor unique identification purposes |
| sampleId | string | | MANDATORY | one sample can have multiple isolates |
| patientId | string | | MANDATORY | reference to Patient.patientId |
| sampleDate | date | YYYY/MM/DD | MANDATORY | |
| sampleWardId | string | | OPTIONAL | Ward where the patient was on day of culture; if on multiple wards, take the first one. |
| isolateNumber | integer | | MANDATORY | If posNeg = 0 (neg), set to 0 |
| bcMicroorgLocalId | string | | MANDATORY | Local ID of the microorganism in lab system (this serves as basis for the algorithm). Should curate this for example if determination for some species is unreliable (e.g. E. cloacae complex, B. cereus complex). Set to “NULL” if posNeg = 0 (neg) |
| posNeg | boolean | 0 (neg); 1 (pos) | MANDATORY | |

Always acquire latest version via PRAISE Network

| | | | | |
|---|---|---|---|---|
| **MOVEMENT.CSV** | | | | This should include all patient movements if it is also used to calculate the denominator[1] |
| movementId | string | | MANDATORY | |
| patientId | string | | MANDATORY | reference to Patient.patientId |
| wardId | string | | MANDATORY | |
| admissionWardDate | date | YYYY/MM/DD | MANDATORY | |
| dischargeWardDate | date | YYYY/MM/DD | MANDATORY | Can be empty if patient is still admitted, i.e. no discharge date available |
| admissionHospDate | date | YYYY/MM/DD | MANDATORY | |
| dischargeHospDate | date | YYYY/MM/DD | MANDATORY | Can be empty if patient is still admitted, i.e. no discharge date available |

[1]**:** Providing all patient movement data can be very voluminous and not easy to acquire. In that situation it may be considered to provide aggregated denominator data as described in Stage 2, complement with movement data of all patients with a blood culture taken

#### Stage 2

| **Variable** | **Data type** | **Values or format (if applicable)** | **Optional?** | **Comments** |
|---|---|---|---|---|
| **PATIENTS.CSV** | | | | |
| patientId | string | | MANDATORY | Should be pseudonymised |
| patientSex | string | M (male); F (female); X (not male not female); U (unknown) | MANDATORY | |
| patientBirthDate | date | YYYY/MM/DD | MANDATORY | For sharing purpose consider dummy |
| patientDeathDate | date | YYYY/MM/DD | OPTIONAL | |
| **BLOODCULTURES.CSV** | | | | *Only positive blood cultures are needed in stage 2* |
| bcId | string | | MANDATORY | A unique identifier of BC microorganism. Example compound sampleId+isolateNumber. Must add isolateNumber for unique identification purposes |
| sampleId | string | | MANDATORY | one sample can have multiple isolates |
| patientId | string | | MANDATORY | reference to Patient.patientId |
| sampleDate | date | YYYY/MM/DD | MANDATORY | |
| sampleWardId | string | | MANDATORY | Ward where culture was taken -> if not present in hospital data then ward where pt was admitted on day of culture; if pt was transferred on day of blood culture, take first ward (where patient came from) |
| isolateNumber | integer | | MANDATORY | |

Always acquire latest version via PRAISE Network

| | | | | |
|---|---|---|---|---|
| bcMicroorgLocalId | string | | MANDATORY | Local ID of the microorganism in lab system = basis for algo. Should curate this for example if determination for some species is unreliable (e.g. E. cloacae complex, B. cereus complex). |
| attributableWardId | string | | MANDATORY | Ward where pt was 2 days prior to culture taken if patient was transferred on day of blood culture, take first ward (where patient came from) |
| admissionHospDate | date | YYYY/MM/DD | MANDATORY | |
| **DENOMINATOR.CSV** | **(aggregated data)** | | | **Denominator (calculated by data provider)** These are denominator data of all patients wardGroup under analysis. See examples below |
| wardGroupType | string | value_set<br>HOSPITAL<br>WARD<br>LOCALWARD<br>ECDCWARD | MANDATORY | Defines at what level the analysis will be done, for example enter hospital, individual wards, local ward groups or ECDCWard groups |
| wardGroupValue | string | value_set containing a value~~s~~ applicable to wardGroups | MANDATORY | Specifies the value options for the denominators. |
| patientDays | numeric | | MANDATORY | Number of patient days in wardGroupValue within specified period type. Midnight method: assign patient day to ward where patient was at 00:00 |
| periodType | value_set | Valueset<br>DAY<br>MONTH<br>QUARTER<br>YEAR | MANDATORY | If possible, DAY, else the lowest level feasible. |
| calendarDateStart | date | YYYY/MM/DD | MANDATORY | Start date to which the denominator value applies. If Period type is DAY, then calendarDateStart = calendarDateEnd |
| calendarDateEnd | Date | YYYY/MM/DD | MANDATORY | end date to which the denominator value applies (inclusive). If Period type is DAY, then calendarDateStart = calendarDateEnd |
| numberOfAdmissions | numeric | | MANDATORY | Number of admissions to that specific WardGroupValue on specified period type |
| numberOfBloodCultureSamples | numeric | | MANDATORY | Count distinct sampleID If available, else try to submit all BC's according to stage 1 (sampleWard, also for negative cultures) |

*See examples of denominator calculation below.*

Always acquire latest version via PRAIS[illegible]

#### Look-up tables

| |
|---|
| **WARDCLASSIFICATION.CSV** |

| | | | | |
|---|---|---|---|---|
| wardId | string | | MANDATORY | |
| wardECDCWard | string | value_set (ECDC classification list) | MANDATORY | ECDC ward specialty classification of ward |
| wardECDCWardPatient | string | value_set (ECDC classification list) | OPTIONAL | ECDC patient specialty classification of ward |
| wardLocalWardGroup | string | value_set (own ward classification hospital) | OPTIONAL | Local classification of ward. If wards have a 1:n relation with ward groups, the aggregate groups can be calculated from the output. |
| **MICROCLASSIFICATION.CSV** | | | | |
| bcMicroorgLocalId | string | | MANDATORY | Local ID of the microorganism in lab system = basis for algo. Should curate this; for example, if determination for some species is unreliable (e.g. E. cloacae complex, B. cereus complex). |
| bcMicroorgSnomedCTConceptId | string | | OPTIONAL | If SNOMED CT code is not available, leave empty |
| bcMicroorgSpecifiedName | string | Description of the bacterial species | OPTIONAL | SNOMED CT label or local label |
| isCommCommensal | boolean | 0 (no); 1 (yes) | MANDATORY if no SNOMED CT codes | If filled use this value; otherwise refer to CDC Common commensal classification, else treat as pathogen |

#### Microorganism classification (based on CDC Common commensals).

- The algorithm runs on the microorganism ID's in place locally. They may be mapped to the CDC common commensal list by SNOMED CT code or manually.
- Local additions may be necessary, for example for new species or different local terminology.
- Please ensure correct grouping of difficult to distinguish pathogens if appropriate. For example, MALDI-ToF cannot reliably distinguish certain species of the *E. cloacae complex*. Reporting species names may lead to incorrect detection of repeat HOB's or polymicrobial HOB's with different species within this complex. Data providers may ensure that they report all relevant Enterobacter species as *E. cloacae complex.*
- Link to Common commensal list (most current version): master-organism-com-commensals-lists.xlsx (live.com)

#### Ward classification

- Wards are classified by ECDC ward specialty for reporting purposes.
- Data providers/hospitals may also perform analyses by ECDC patientSpecialty or a local ward classification system.
- Link to ECDC classification: European Centre for Disease Prevention and Control. Point prevalence survey of healthcare-associated infections and antimicrobial use in European acute care hospitals – protocol version 6.1. Stockholm: ECDC; 2022.: Point prevalence survey of HAIs and antimicrobial use in European acute care hospitals, protocol version 6.1 (europa.eu)

Always acquire latest version via PRAISE Network

### Minimal dataset (output) of the PRAISE-HOB algorithm

**Notes:**

The output file should at least have one line for each HOB microorganism event (line listing) hence there is more than 1 line per HOB possible (if polymicrobial event);
For debugging purposes, it is possible to allow for one line for each positive blood culture (see hobBcStatus)

Cardinality:

[1 … n]: one or more lines of data per patient
[0 … n]: may be left empty if not applicable or when de data is optional.

| Variable | Data type | Values or format (if applicable) | Cardinality | Notes |
|---|---|---|---|---|
| **HOBOUTPUT.CSV** | | | | |
| hobId | string | | [1..n] | string composed of [patientId]_[hobEventDate in yyyyMMDD], so 1022_20220122 |
| hobEventDate | date | YYYY/MM/DD | [1..n] | Date of the HOB (first positive BC finding within HOB episode) |
| patientId | string | | [1..n] | |
| patientSex | string | M (male); F (female); X (not male not female); U (unknown) | [1..n] | |
| patientBirthDate | date | YYYY/MM/DD | [1..n] | |
| patientDeathDate | date | YYYY/MM/DD | [0..n] | |
| bcId | string | | [1..n] | for example sampleId+IsolateNumber //=bloodCultureId |
| bcSampleDate | date | YYYY/MM/DD | [1..n] | |
| hobBcStatus | value_set | **MICROORG_EVENT**; COPY_STRAIN; CC_CONF; SOLIT_COMM | [1..n] | Value MICROORG_EVENT mandatory, other categories are optional but may be assigned for debugging purposes |
| bcMicroorgLocalId | string | | [1..n] | |
| bcMicroorgSnomedCTConceptId | string | value_set | [0..n] or [1…n] if SNOMED CT | MANDATORY if SNOMED CT implemented, 1 microorganism per line |
| bcMicroorgSpecifiedName | string | label with belonging to SNOMED CT or localId | [0..n] | |
| bcIsCommCommensal | boolean | 0 (no); 1 (yes) | [1..n] | |
| hobIsPolymicrobial | boolean | 0 (no); 1 (yes) | [1..n] | if polymicrobial -> 1 = True = Yes |
| hobWithCommCommensal | boolean | 0 (no); 1 (yes) | [1..n] | HOB contains a CSC |

Always acquire latest version via PRAISE-Network

| | | | | |
|---|---|---|---|---|
| hobWithPathogen | boolean | 0 (no); 1 (yes) | [1..n] | HOB contains a recognized pathogen |
| hobAttrWardId | string | | [1..n] | WardID to which HOB is attributed |
| hobAttrWardECDCWard | string | value_set (ECDC classification list) | [1..n] | Ward type to what HOB is attributed: Consensus = ward type, patient specialty = optional. |
| hobAttrWardECDCWardPatient | string | value_set (ECDC classification list) | [0..n] | |
| hobAttrWardLocalWardGroup | string | value_set (locally defined) | [0..n] | |
| **DENOMINATOR.CSV** | **(aggregated data)** | | | **Denominator (intell in hospital/facility)** These are denominator data of all patients in the ward. |
| wardGroupType | string | value_set<br>HOSPITAL<br>WARD<br>LOCALWARD<br>ECDCWARD | MANDATORY | Defines at what level the analysis will be done, for example wardID, ECDCWard, or other local classifications |
| wardGroupValue | string | value_set containing a values applicable to wardGroups | MANDATORY | Specifies the value options for the denominators. |
| patientDays | numeric | | MANDATORY | Number of patient days in wardGroupValue within specified period type.<br>Midnight method: assign patient day to ward where patient was at 00:00 |
| periodType | value_set | Value_set<br>DAY<br>MONTH<br>QUARTER<br>YEAR | MANDATORY | If possible DAY, else the lowest level feasible. |
| calendarDateStart | date | YYYY/MM/DD | MANDATORY | Start date to which the denominator value applies. If Period type is DAY, then calendarDateStart = calendarDateEnd |
| calendarDateEnd | Date | YYYY/MM/DD | MANDATORY | End date to which the denominator value applies (inclusive). If Period type is DAY, then calendarDateStart = calendarDateEnd |
| numberOfAdmissions | numeric | | MANDATORY | Number of admissions to that specific WardGroupValue on specified period type |
| numberOfBloodCultureSamples | numeric | | MANDATORY | Count distinct sampleID If available, else try to submit all BC's according to stage 1 (sampleWard, also for negative cultures) |

Always acquire latest version via PRAISE Netw

### Examples for denominator calculation

#### Example of the midnight method for use of patient days as denominator

| Hospital admission day | 0 | 1 | 2 | 3 | 4 | 5 | 6 | 7 | 8 | 9 | 10 |
|---|---|---|---|---|---|---|---|---|---|---|---|
| Ward name | Ward 1<br>*Admission to ward 1 at 8:00* | Ward 1 | Ward 1 | Ward 1 | Ward 1<br>*Transfer to Ward 2 at 10:00* | Ward 2 | Ward 2 | Ward 2<br>*Transfer to Ward 3 at 23:55* | Ward 3 | Ward 3 | Ward 3<br>*Hospital discharge at 15:00* |
| Ward specialty | ICU | ICU | ICU | ICU | ICU | MED | MED | MED | SUR | SUR | SUR |
| Count in denominator | none | ICU | ICU | ICU | ICU | MED | MED | MED | SUR | SUR | SUR |

Notes: Day 0 is not counted in the denominator data because the patient was not yet in the ward at midnight (00:00), Day 4 is counted as ICU since the patient was in ICU at midnight.

#### Examples of denominator data analysis by wardGroupType and wardGroupValue

In the denominator data, different ward classification methods are included as possibilities to report HOB rates, hence giving different units of analysis. The choice of reporting unit may differ by purpose (e.g., sharing with external institution for trend monitoring vs. in-hospital reporting).

For each unit within the period type chosen (day, month, quarter, year), the denominator count is report per unit of analysis. And multiple aggregation levels may be included in one dataset.

This approach allows for flexible aggregation of output data.

In the following example, the hospital has 6 wards with different specialties, and each ward has 4 beds, and we assume they are always occupied.

The Look-up table may have this structure:

| Ward name | Description | wardECDCWard | wardECDCWardPatient | wardLocalWardGroup |
|---|---|---|---|---|
| A1 | ICU medical 1 | ICU | ICUMED | ICU |
| B2 | Internal A | MED | MEDHEMA | Medical tower |
| C3 | Internal B | MED | MEDNEPH | Medical tower |
| D4 | Neurology & neurosurgery | MIX | MIX | NEURO |
| E5 | General surgery | SUR | SURGEN | Surgical tower |
| F6 | Orthopaedic | SUR | SURGEN | Surgical Stepdown |

Always acquire latest version via PRAISE Network

**Denominator data for the whole hospital** (period type = DAY)

| wardGroupType | wardGroupValue | patientDays | periodType | calendarDateStart | calendarDateEnd | numberOfWardGroupAdmissions | numberOfBloodCultureSamples |
|---|---|---|---|---|---|---|---|
| HOSPITAL | HOSP | 24 | DAY | 2023/01/01 | 2023/01/01 | 5 | 10 |
| HOSPITAL | HOSP | 24 | DAY | 2023/01/02 | 2023/01/02 | 3 | 20 |
| HOSPITAL | HOSP | 24 | DAY | 2023/01/03 | 2023/01/03 | 5 | 15 |
| HOSPITAL | HOSP | 24 | DAY | 2023/01/04 | 2023/01/04 | 2 | 35 |

**Denominator data by ECDCWard level grouping** (period type = DAY)

| wardGroupType | wardGroupValue | patientDays | periodType | calendarDateStart | calendarDateEnd | numberOfWardGroupAdmissions | numberOfBloodCultureSamples |
|---|---|---|---|---|---|---|---|
| ECDCWARD | MED | 8 | DAY | 2023/01/01 | 2023/01/01 | 1 | 2 |
| ECDCWARD | MED | 8 | DAY | 2023/01/02 | 2023/01/02 | 0 | 5 |
| ECDCWARD | MED | 8 | DAY | 2023/01/03 | 2023/01/03 | 1 | 3 |
| ECDCWARD | MED | 8 | DAY | 2023/01/04 | 2023/01/04 | 0 | 8 |
| ECDCWARD | SUR | 8 | DAY | 2023/01/01 | 2023/01/01 | 0 | 2 |
| ECDCWARD | SUR | 8 | DAY | 2023/01/02 | 2023/01/02 | 1 | 5 |
| ECDCWARD | SUR | 8 | DAY | 2023/01/03 | 2023/01/03 | 1 | 3 |
| ECDCWARD | SUR | 8 | DAY | 2023/01/04 | 2023/01/04 | 0 | 8 |
| ECDCWARD | MIX | 4 | DAY | 2023/01/01 | 2023/01/01 | 2 | 3 |
| ECDCWARD | MIX | 4 | DAY | 2023/01/02 | 2023/01/02 | 1 | 5 |
| ECDCWARD | MIX | 4 | DAY | 2023/01/03 | 2023/01/03 | 2 | 3 |
| ECDCWARD | MIX | 4 | DAY | 2023/01/04 | 2023/01/04 | 1 | 10 |
| ECDCWARD | ICU | 4 | DAY | 2023/01/01 | 2023/01/01 | 2 | 3 |
| ECDCWARD | ICU | 4 | DAY | 2023/01/02 | 2023/01/02 | 0 | 5 |
| ECDCWARD | ICU | 4 | DAY | 2023/01/03 | 2023/01/03 | 1 | 6 |
| ECDCWARD | ICU | 4 | DAY | 2023/01/04 | 2023/01/04 | 1 | 9 |

Always acquire latest version via PRAISE Network

**Denominator data by Local grouping** (period type = DAY)

| wardGroupType | wardGroupValue | patientDays | periodType | calendarDateStart | calendarDateEnd | numberOfWardGroupAdmissions | numberOfBloodCultureSamples |
|---|---|---|---|---|---|---|---|
| LOCALWARD | Medical tower | 8 | DAY | 2023/01/01 | 2023/01/01 | 1 | 2 |
| LOCALWARD | Medical tower | 8 | DAY | 2023/01/02 | 2023/01/02 | 0 | 5 |
| LOCALWARD | Medical tower | 8 | DAY | 2023/01/03 | 2023/01/03 | 1 | 3 |
| LOCALWARD | Medical tower | 8 | DAY | 2023/01/04 | 2023/01/04 | 0 | 8 |
| LOCALWARD | Surgical tower | 4 | DAY | 2023/01/01 | 2023/01/01 | 0 | 1 |
| LOCALWARD | Surgical tower | 4 | DAY | 2023/01/02 | 2023/01/02 | 1 | 3 |
| LOCALWARD | Surgical tower | 4 | DAY | 2023/01/03 | 2023/01/03 | 0 | 1 |
| LOCALWARD | Surgical tower | 4 | DAY | 2023/01/04 | 2023/01/04 | 0 | 4 |
| LOCALWARD | Surgical stepdown | 4 | DAY | 2023/01/01 | 2023/01/01 | 0 | 1 |
| LOCALWARD | Surgical stepdown | 4 | DAY | 2023/01/02 | 2023/01/02 | 0 | 2 |
| LOCALWARD | Surgical stepdown | 4 | DAY | 2023/01/03 | 2023/01/03 | 1 | 2 |
| LOCALWARD | Surgical stepdown | 4 | DAY | 2023/01/04 | 2023/01/04 | 0 | 4 |
| LOCALWARD | NEURO | 4 | DAY | 2023/01/01 | 2023/01/01 | 2 | 3 |
| LOCALWARD | NEURO | 4 | DAY | 2023/01/02 | 2023/01/02 | 1 | 5 |
| LOCALWARD | NEURO | 4 | DAY | 2023/01/03 | 2023/01/03 | 2 | 3 |
| LOCALWARD | NEURO | 4 | DAY | 2023/01/04 | 2023/01/04 | 1 | 10 |
| LOCALWARD | ICU | 4 | DAY | 2023/01/01 | 2023/01/01 | 2 | 3 |
| LOCALWARD | ICU | 4 | DAY | 2023/01/02 | 2023/01/02 | 0 | 5 |
| LOCALWARD | ICU | 4 | DAY | 2023/01/03 | 2023/01/03 | 1 | 6 |
| LOCALWARD | ICU | 4 | DAY | 2023/01/04 | 2023/01/04 | 1 | 9 |

Always acquire latest version via PRAISE Network

**Denominator data by single ward** (period type = DAY)

| wardGroupType | wardGroupValue | patientDays | periodType | calendarDateStart | calendarDateEnd | numberOfWardGroupAdmissions | numberOfBloodCultureSamples |
|---|---|---|---|---|---|---|---|
| WARD | B2 | 4 | DAY | 2023/01/01 | 2023/01/01 | 1 | 1 |
| WARD | B2 | 4 | DAY | 2023/01/02 | 2023/01/02 | 0 | 2 |
| WARD | B2 | 4 | DAY | 2023/01/03 | 2023/01/03 | 0 | 1 |
| WARD | B2 | 4 | DAY | 2023/01/04 | 2023/01/04 | 0 | 4 |
| WARD | C3 | 4 | DAY | 2023/01/01 | 2023/01/01 | 0 | 1 |
| WARD | C3 | 4 | DAY | 2023/01/02 | 2023/01/02 | 0 | 3 |
| WARD | C3 | 4 | DAY | 2023/01/03 | 2023/01/03 | 1 | 2 |
| WARD | C3 | 4 | DAY | 2023/01/04 | 2023/01/04 | 0 | 4 |
| WARD | E5 | 4 | DAY | 2023/01/01 | 2023/01/01 | 0 | 1 |
| WARD | E5 | 4 | DAY | 2023/01/02 | 2023/01/02 | 1 | 3 |
| WARD | E5 | 4 | DAY | 2023/01/03 | 2023/01/03 | 0 | 1 |
| WARD | E5 | 4 | DAY | 2023/01/04 | 2023/01/04 | 0 | 4 |
| WARD | F6 | 4 | DAY | 2023/01/01 | 2023/01/01 | 0 | 1 |
| WARD | F6 | 4 | DAY | 2023/01/02 | 2023/01/02 | 0 | 2 |
| WARD | F6 | 4 | DAY | 2023/01/03 | 2023/01/03 | 1 | 2 |
| WARD | F6 | 4 | DAY | 2023/01/04 | 2023/01/04 | 0 | 4 |
| WARD | D4 | 4 | DAY | 2023/01/01 | 2023/01/01 | 2 | 3 |
| WARD | D4 | 4 | DAY | 2023/01/02 | 2023/01/02 | 1 | 5 |
| WARD | D4 | 4 | DAY | 2023/01/03 | 2023/01/03 | 2 | 3 |
| WARD | D4 | 4 | DAY | 2023/01/04 | 2023/01/04 | 1 | 10 |
| WARD | A1 | 4 | DAY | 2023/01/01 | 2023/01/01 | 2 | 3 |
| WARD | A1 | 4 | DAY | 2023/01/02 | 2023/01/02 | 0 | 5 |
| WARD | A1 | 4 | DAY | 2023/01/03 | 2023/01/03 | 1 | 6 |
| WARD | A1 | 4 | DAY | 2023/01/04 | 2023/01/04 | 1 | 9 |

Always acquire latest version via PRAISE Network
